# Predilection for Perplexion: Preoperative microstructural damage is linked to postoperative delirium

**DOI:** 10.1101/2025.01.08.24319243

**Authors:** Tyler H. Reekes, Vinith R. Upadhya, Jenna L. Merenstein, Mary Cooter-Wright, David J. Madden, Melody A. Reese, Piper C. Boykin, Noah J. Timko, Judd W. Moul, Grant E. Garrigues, Katherine T. Martucci, Harvey Jay Cohen, Heather E. Whitson, Joseph P. Mathew, Michael J. Devinney, Henrik Zetterberg, Kaj Blennow, Leslie M. Shaw, Teresa Waligorska, Jeffrey N. Browndyke, Miles Berger, MADCO-PC and INTUIT Investigators

**Affiliations:** Department of Anesthesiology, Duke University Medical Center, Durham, NC; Trinity College of Arts and Sciences, Duke University, Durham, NC; Brain Imaging and Analysis Center, Duke University Medical Center, Durham, NC; Department of Psychiatry and Behavioral Sciences, Duke University Medical Center, Durham, NC; Department of Anesthesiology, Perioperative and Pain Medicine, Stanford University School of Medicine, Stanford, CA; Department of Surgery, Duke University Medical Center, Durham, NC; Midwest Orthopedics at Rush, Rush University, Chicago, IL; Center for the Study of Aging and Human Development, Duke University Medical Center, Durham, NC; Department of Psychiatry and Neurochemistry, Institute of Neuroscience and Physiology, Sahlgrenska Academy, University of Gothenburg, Mölndal, Sweden; Department of Neurodegenerative Disease, UCL Institute of Neurology, Queen Square, London, UK; Clinical Neurochemistry Laboratory, Sahlgrenska University Hospital, Mölndal, Sweden; UK Dementia Research Institute at UCL, London, UK; Hong Kong Center for Neurodegenerative Diseases, Clear Water Bay, Hong Kong, China; Wisconsin Alzheimer’s Disease Research Center, University of Wisconsin School of Medicine and Public Health, University of Wisconsin-Madison, Madison, WI, USA; Department of Pathology and Laboratory Medicine, University of Pennsylvania Perelman School of Medicine, Philadelphia, PA; Duke Institute for Brain Sciences, Duke University, Durham, NC

**Keywords:** Postoperative delirium, diffusion-weighted imaging, attention, neurite density, atrophy

## Abstract

Postoperative delirium is the most common postsurgical complication in older adults and is associated with an increased risk of long-term cognitive decline and Alzheimer’s disease (AD) and related dementias (ADRD). However, the neurological basis of this increased risk— whether postoperative delirium unmasks latent preoperative pathology or leads to AD-relevant pathology after perioperative brain injury—remains unclear. Recent advancements in neuroimaging techniques now enable the detection of subtle brain features or damage that may underlie clinical symptoms. Among these, Neurite Orientation Dispersion and Density Imaging (NODDI) can help identify microstructural brain damage, even in the absence of visible macro-anatomical abnormalities.

To investigate potential brain microstructural abnormalities associated with postoperative delirium and cognitive function, we analyzed pre- and post-operative diffusion MRI data from 111 patients aged ≥60 years who underwent non-cardiac/non-intracranial surgery. Specifically, we investigated preoperative variation in diffusion metrics within the posterior cingulate cortex (PCC), a region in which prior work has identified glucose metabolism alterations in the delirious brain, and a key region in the early accumulation of amyloid beta (Aβ) in preclinical AD. We also examined the relationship of preoperative PCC NODDI abnormalities with preoperative cognitive function. Compared to patients who did not develop postoperative delirium (n=99), we found increased free water (FISO) and neurite density index (NDI) and decreased orientation dispersion index (ODI) in the dorsal PCC before surgery among those who later developed postoperative delirium (n=12). These FISO differences before surgery remained present at six weeks postoperatively, while these NDI and ODI differences did not. Preoperative dorsal PCC NDI and ODI values were also positively associated with preoperative attention/concentration performance, independent of age, education level, and global brain atrophy. Yet, these diffusion metrics were not correlated with cerebrospinal fluid Aβ positivity or levels.

These results suggest that preoperative latent brain abnormalities within the dorsal PCC may underlie susceptibility to postoperative delirium, independent of AD-related (i.e., Aβ) neuropathology. Furthermore, these preoperative microstructural differences in the dorsal PCC were linked to preoperative deficits in attention/concentration, a core feature of postoperative delirium. Our findings highlight microstructural vulnerability within the PCC, a key region of the default mode network, as a neuroanatomic locus that can help explain the link between preoperative attention/concentration deficits and increased postoperative delirium risk among vulnerable older surgical patients.

## Introduction

Postoperative delirium is the most common postsurgical complication among adults over the age of 65.^1^ Previous estimates suggest that each year, >19 million older Americans undergo surgery.^2^ Postoperative delirium is characterized by fluctuating changes in the levels of consciousness and attention, and is associated with increased hospital length of stay, nursing home placement, and mortality,^3–5^ and an increased risk for Alzheimer’s disease (AD) and related dementias (ADRD).^6^

The etiology and pathophysiology of delirium remain unclear, yet several factors are associated with increased postoperative delirium risk, such as preexisting cognitive impairment^7^ or dementia, increased age, and greater co-morbidity burden.^8^ Yet, these risk factors are neither highly sensitive nor specific for delirium, and the neurologic or neuroanatomic correlates of these factors have remained unknown. However, prior studies have revealed altered functional connectivity (measured via functional magnetic resonance imaging [fMRI]) and hypometabolism (measured via positron emission tomography [PET]) in the posterior cingulate cortex (PCC) in patients with postoperative cognitive dysfunction and delirium, respectively.^9–16^ The PCC is also a brain region in which amyloid beta (Aβ) accumulates early in preclinical AD,^17^ and there are epidemiologic links between postoperative delirium and ADRD. Thus, we hypothesized that gray matter microstructural abnormalities within the PCC would be associated with preoperative cognitive impairment, postoperative delirium risk, and Aβ pathology. To test these hypotheses, we examined diffusion MRI data among older surgical patients and utilized the biophysical multicompartment <u>N</u>eurite <u>O</u>rientation <u>D</u>ispersion and <u>D</u>ensity <u>I</u>maging (NODDI) model to estimate the microstructural composition of brain tissue.^18^ We compared preoperative diffusion metrics within the PCC among patients who later developed (versus those who did not develop) postoperative delirium. Next, we measured the association between PCC diffusion metrics and several measures of baseline cognitive function. Lastly, we compared preoperative diffusion metrics within the PCC to preoperative cerebrospinal fluid (CSF) Aβ levels to determine the extent to which ADRD pathology may be associated with preoperative PCC microstructural abnormalities.

## Materials and Methods

### Participants

Data were obtained from two previously conducted observational, prospective cohort studies, MADCO-PC [clinicaltrials.gov NCT01993836] and INTUIT [clincaltrials.gov NCT03273335], both of which were approved by the Duke University Health System institutional review board prior to study initiation. Informed consent was obtained from all subjects prior to study participation in accordance with the Declaration of Helsinki. Inclusion criteria for both MADCO-PC and INTUIT studies were age ≥ 60 years, English fluency, and a home address within a ∼1 hour drive of Duke University Medical Center. Exclusion criteria included incarceration, pregnancy, and/or active treatment with chemotherapy drugs with known cognitive side effects. INTUIT also excluded patients on immunomodulatory drugs at the time of surgery, since these could potentially interfere with the neuroinflammatory markers under study in INTUIT.^19^ Participants were excluded from MRI data collection if they were unsafe to scan (e.g., those with MRI-incompatible implants/devices).

### Delirium Assessment

MADCO-PC participants were screened once daily for postoperative delirium using the Confusion Assessment Method (CAM)^20^ until hospital discharge. INTUIT participants were screened twice daily for delirium via the 3-Minute Diagnostic Confusion Assessment Method (3D-CAM)^21^ or the Confusion Assessment Method for the Intensive Care Unit (CAM-ICU; used for intubated patients)^22^ on postoperative days 1-5 or until hospital discharge. These assessments were completed by study technicians trained by a licensed clinical neuropsychologist (JNB). Delirium assessors were trained with materials from the Hebrew SeniorLife Program/Harvard Medical School SAGES study group.^23^ Assessors performed delirium assessments on study patients only after achieving a ≥ 90% agreement rate with standardized training assessments. The accuracy of our assessors, as measured by these standardized video-taped assessments, was 93%.^23^ All assessors also continually received feedback to further enhance accuracy. Additionally, since the fluctuating nature of delirium means that it can be missed by assessments at single time points, delirium was also assessed via a validated chart review method.^24^ Participants with one or more positive CAM, 3D-CAM, or CAM-ICU assessments or a positive delirium chart review were considered to be positive for postoperative delirium.

### Cognitive Assessment

The neurocognitive battery^19,25^ included standardized assessments of auditory-verbal memory, immediate and delayed recall of unstructured narrative and structured word-list stimuli, information processing speed and executive function (i.e., logical sequencing and task switching), immediate and delayed recall of visual memory for simple line drawing stimuli, and auditory-verbal simple and complex attention (i.e., working memory). From study participants who had complete baseline cognitive test scores from the parent MADCO-PC and INTUIT studies, a factor analysis with oblique rotation was performed on the 14 cognitive variables in the neurocognitive battery.^26^ The number of factors was selected to explain at least 80% of the variance in cognitive test scores. This identified a five-factor solution that explained 82% of the variance in the original test scores, corresponding to five cognitive domains: structured auditory-verbal memory (ability to recall from a structured word list), unstructured auditory-verbal memory (ability to remember from a narrative), visual memory, information processing speed/executive functioning, and auditory-verbal attention/concentration (individual tests and scores presented in supplementary Table 1). Additionally, the average of the five cognitive domain scores was used as a summary metric for global cognitive performance, i.e., the continuous cognitive index (CCI), as described in multiple prior publications from our group.^25,27–29^

### Imaging Data

#### Acquisition

Imaging data were acquired at the Duke Brain Imaging and Analysis Center (BIAC) on a 3T GE Signa MR750 MRI scanner (GE Healthcare, Waukesha, WI, USA) equipped with an 8 channel receive-only head coil. Participants wore earplugs to reduce scanner noise; foam pads were used to minimize head motion.

Anatomical data consisted of a high-resolution T1-weighted fast spoiled gradient-echo image volume (FSPGR) with the following parameters: oblique axial acquisition, TR 6.93 ms, TE 3.0 ms, 11° flip angle, 1 mm slice thickness, 256 × 256 mm matrix, and 256 x 256 mm field of view (FOV). Whole-brain diffusion-weighted imaging (DWI) data were acquired using a single-shot echo planar imaging (EPI) sequence with the following parameters: anterior to posterior acquisition, TR = 17 ms, TE = 80.8 ms, flip angle = 90°, 2 mm slice thickness and spacing between slices = 2 mm, 128 x 128 matrix, with a 256 x 256 reconstructed matrix. Diffusion-weighted gradients were applied in 25 directions with *b* values of 1000 s/mm^2^ with two non-diffusion-weighted *b* = 0 images. A reverse phase sequence was not obtained.

#### Processing

Raw DWI data were preprocessed using MRtrix3^30^ and FSL (FMRIB Software Library)^31^ as previously described.^32^ The data were denoised (*dwidenoise*) and corrected for motion and eddy current-induced distortions (*dwifslpreproc*). Lastly, all data were bias-corrected (*dwibiascorrect*), and non-brain tissue was removed to generate a whole-brain mask (*dwi2mask*). All processed DWI scans were visually inspected for artifacts and anatomical abnormalities by trained researchers (THR, VRU). The processed DWI data were then used as inputs to the NODDI MATLAB toolbox (https://www.nitrc.org/projects/noddi_toolbox) to obtain diffusion estimates.^33^ Diffusion estimates derived from NODDI include the (1) volume fraction of isotropic diffusion (FISO), (2) neurite density index (NDI), and (3) orientation dispersion index (ODI; see supplementary Figure 1). FISO measures the fraction of a voxel occupied by isotropic (free) water that can diffuse equally in all directions. NDI represents the fraction of a voxel occupied by neuronal cell bodies and processes (e.g., dendrites and axons). ODI quantifies the variability in the orientation of neuronal processes within a voxel, reflecting the complexity and angular dispersion of the extracellular space around these processes.^34–36^

#### Analysis

To obtain diffusion estimates from specific brain regions, we used Gray Matter-Based Spatial Statistics (GBSS),^37,38^ which adopts a similar framework to FSL’s Tract Based Spatial Statistics (TBSS).^39^ This voxel-wise approach skeletonizes the gray matter and projects diffusion metrics from the most probable local gray matter voxel onto the skeleton to facilitate subsequent comparisons between groups.

First, for each participant, we estimated the fraction of CSF based on isotropic diffusion maps and the fraction of white matter or non-white matter tissue based on fractional anisotropy maps. The fraction of gray matter tissue was then obtained by subtracting the fraction of CSF and white matter from 1 in each voxel. To improve tissue contrast and between-subject registration, we multiplied each partial volume estimation map by their corresponding contrast (0 = CSF, 1 = gray matter, 2 = white matter) and summed them together to create an image similar to a structural T1-weighted image in which there is a distinct delineation between tissue types. The resulting images were then used for groupwise nonlinear registration into Montreal Neurological Institute (MNI152)^40^ space with 1 x 1 x 1 mm resolution using the Advanced Normalization Tools *buildtemplateparallel.sh* script.^41^

For each participant, the gray matter fraction, NDI, and ODI maps were warped to MNI152 space using the warp fields estimated by the Advanced Normalization Tools *buildtemplateparallel.sh* script. The skeleton was obtained by averaging the aligned gray matter probability maps across participants and thinning the skeleton so that it only contained voxels with a high probability of being gray matter (i.e., gray matter fraction probability > 0.65 in > 75% of participants). The remaining voxels that did not meet these criteria were filled with the average diffusion metrics of the surrounding satisfactory voxels on the skeleton, weighted by their closeness with a Gaussian kernel (σ = 2 mm). That is, values from nearby reliable areas were used to estimate information from voxels that did not meet these reliability criteria, with closer areas contributing more to the final value than those further away. The aligned FISO, NDI, and ODI maps were then projected onto the mean gray matter skeleton for each participant. Lastly, to limit analyses to the key region of interest, we multiplied the mean gray matter skeleton by a binarized standard MNI defined PCC mask (Harvard-Oxford cortical atlas with 50% probability and 1 x 1 x 1 mm resolution ^42,43^).

### CSF Aβ measurements

CSF samples were obtained as previously described.^19,44^ CSF levels of the 42 amino acid-form of Aβ (Aβ42) was measured using the INNO-BIA AlzBio3 platform [Innogenetics; Ghent, Belgium]^45^ on samples from 47 of the 50 MADCO-PC subjects, and the Roche Elecsys platform [Roche Diagnostics; Basel, Switzerland]^19^ on samples from 53 of the 61 INTUIT subjects. Aβ42 concentrations were compared against previously described cutoffs for each of these respective assays.^46,47^

### Statistical Analysis

Significant group differences in diffusion metrics were assessed by a voxel-wise analysis limited to the PCC region of interest with threshold-free cluster enhancement (TFCE), via the use of *Randomise* (FSL’s tool for nonparametric permutation inference) with 5,000 permutations. TFCE signal detection is based on both the intensity and spatial extent of contiguous clusters without relying on traditional voxel and cluster thresholds and is thus more sensitive to meaningful neuroanatomical patterns.^48^ For our primary analyses, PCC region of interest constrained TFCE images were generated and thresholded (corrected p < 0.05) to identify contrasts that contained significant voxels at both pre- and postoperative timepoints. For exploratory secondary analyses on the data acquired 6-weeks after surgery, we relaxed the statistical threshold to corrected p < 0.08 to capture any trend-level effects. To determine the subregion localization of voxel subclusters within the PCC (i.e., ventral versus dorsal), we superimposed Brodmann’s areas. We then extracted each average diffusion metric within significant voxels (identified by a contrast between patients who developed versus those did not develop delirium) within the PCC region of interest using *fslstats*.

To better understand significant relationships between baseline diffusion metrics and cognitive factor scores, we then used multivariable linear regression models adjusted for age, education, and global brain atrophy (ventricle to brain ratio), because of their known associations with cognitive performance,^49^ and to control for the potential parallel influence of significant brain volume loss^50^ on diffusion metrics. Relationships between baseline diffusion metrics and cognitive factors or CSF Aβ were assessed via Spearman correlation tests. Analysis of numeric data for CSF Aβ was restricted to the data acquired from the Roche Elecsys platform, due to superior analytical performance (i.e., reduced variability and high reproducibility). We report scaled beta coefficients for diffusion metrics for a 1-unit SD change interpretation. Finally, to assess the stability of the linear regression coefficients and mitigate the impact of outliers, we performed bootstrapping with 1,000 resamples. This non-parametric method creates confidence intervals by repeatedly resampling the data with replacement and refitting the model for each sample.

## Results

### Sample Characteristics

A total of 140 patients were enrolled in the MADCO-PC study and 196 patients in the INTUIT study. Each study included a nested imaging cohort, comprised of 77 and 86 patients respectively, resulting in 163 participants with perioperative imaging. We analyzed MRI data from 113 subjects who had both preoperative and 6-week postoperative high quality (i.e., acceptable motion, and void of artifacts or gross anatomical abnormalities) structural and DWI scans. Of these, 111 had analyzable diffusion MRI and 109 had available baseline cognitive testing data (Figure 1), of whom 12 developed postoperative delirium.

**Figure 1:**
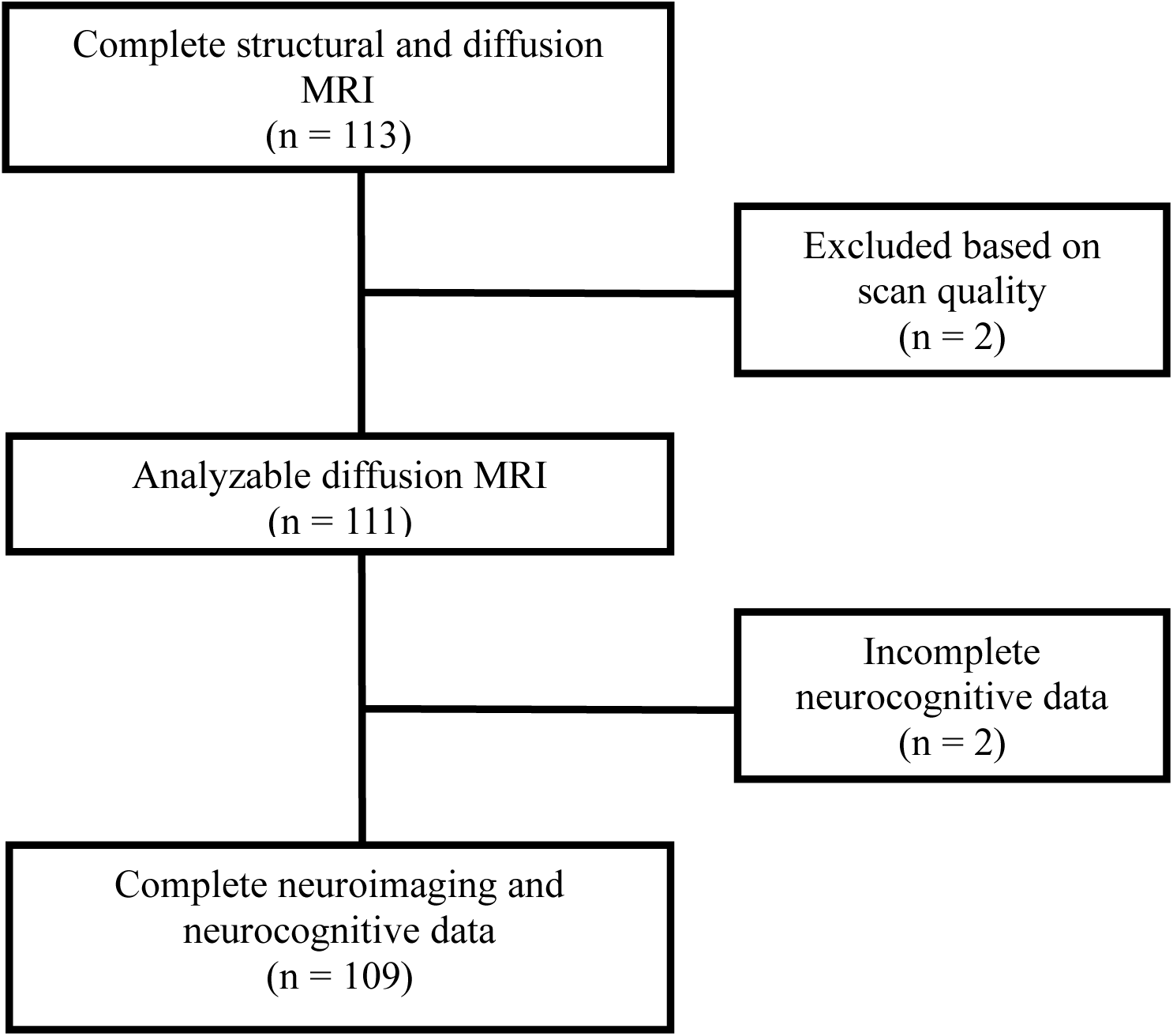
STROBE Diagram

Consistent with prior studies,^7^ patients who developed (versus those who did not develop) postoperative delirium were slightly older (mean (SD): 71.42 (9.01) vs 67.94 (5.13) years); had fewer years of education (14.08 (3.73) vs 15.60 (3.19) years); and lower baseline cognitive function, evidenced by both mini mental state exam (MMSE: 25.58 (3.99) vs 28.41 (1.53) and continuous cognitive index scores (CCI: -0.83 (0.91) vs 0.16 (0.60)) (Table 1). Perioperative factors (Supplementary Table 2) including surgical service, intraoperative age adjusted minimum alveolar concentration (aaMAC), ketamine dose, dexmedetomidine dose, opioid dose (expressed in oral morphine equivalents), muscle relaxant dose (expressed in rocuronium dose equivalents),^51^ and estimated blood loss were similar across groups. Surgical time, however, was longer in those subjects who went on to develop postoperative delirium.

**Table 1.** – Participant Demographics

|  | **Overall** | **No Delirium** | **Delirium** |
| --- | --- | --- | --- |
| **N** | 111 | 99 | 12 |
| **Age (years)** | 68.32 (5.72) | 67.94 (5.13) | 71.42 (9.01) |
| **Sex (%)** |  |  |  |
| Female | 50 (45.0%) | 46 (46.5%) | 4 (33.3%) |
| Male | 61 (55.0%) | 53 (53.5%) | 8 (66.7%) |
| **Race (%)** |  |  |  |
| Black or African American | 14 (12.6%) | 12 (12.1%) | 2 (16.7%) |
| Caucasian/White | 96 (86.5%) | 87 (87.9%) | 9 (75.0%) |
| Other | 1 (0.9%) | 0 (0.0%) | 1 (8.3%) |
| **Education (years) [Q1, Q3]** | 16 [12, 17] | 16 [12.5, 17] | 15 [12, 16.5] |
| **MMSE [Q1, Q3]** | 29 [27.5, 30] | 29 [28, 30] | 27.5 [22, 29] |
| **Height (cm)** | 170.49 (10.44) | 170.86 (10.5) | 167.49 (9.87) |
| **Weight (kg)** | 86.85 (17.94) | 88.14 (17.77) | 76.19 (16.31) |
| **BMI (kg/m<sup>2</sup>)** | 29.91 (5.47) | 30.25 (5.57) | 27.12 (3.67) |
| **VBR (%)** | 2.77 (1.57%) | 2.61 (1.2%) | 4.09 (3.15%) |
| **ASA [Q1, Q3]** | 3.00 [2.00, 3.00] | 3.00 [2.00, 3.00] | 3.00 [3.00, 3.00] |
| **APOE4 carrier (%)** | 37 (33.6%) | 33 (33.7%) | 4 (33.3%) |

Values are presented as either mean (SD), median [Q1, Q3] or number (percentage)
MMSE – Mini Mental State Exam
BMI – Body Mass Index
VBR – Ventricle to Brain Ratio (sum of lateral, 3<sup>rd</sup> and 4<sup>th</sup> ventricles / total parenchymal volume)
ASA – American Society of Anesthesiologists physical status classification system
APOE – Apolipoprotein E

To test whether preoperative gray matter microstructural differences within the PCC might predispose patients to delirium, we used between-subject voxelwise t-tests to assess diffusion metrics within a PCC region of interest between those who later developed postoperative delirium (n = 12) versus those who did not (n = 99). In patients who later developed (vs those who did not develop) postoperative delirium, we found increased preoperative FISO and decreased preoperative NDI and ODI, primarily within the dorsal PCC (Brodmann’s Area d23) (Figure 2; see Table 2 for cluster coordinates). Importantly, the FISO contrast (but not the NDI or ODI contrasts) continued to demonstrate significant differences between groups at 6 weeks after surgery. At a relaxed, more exploratory threshold of p < 0.08, all three diffusion metrics demonstrated between group differences at 6 weeks after surgery, consistent with baseline findings (see Supplementary Figure 2; see Supplementary Table 3 for cluster coordinates).

**Figure 2:**
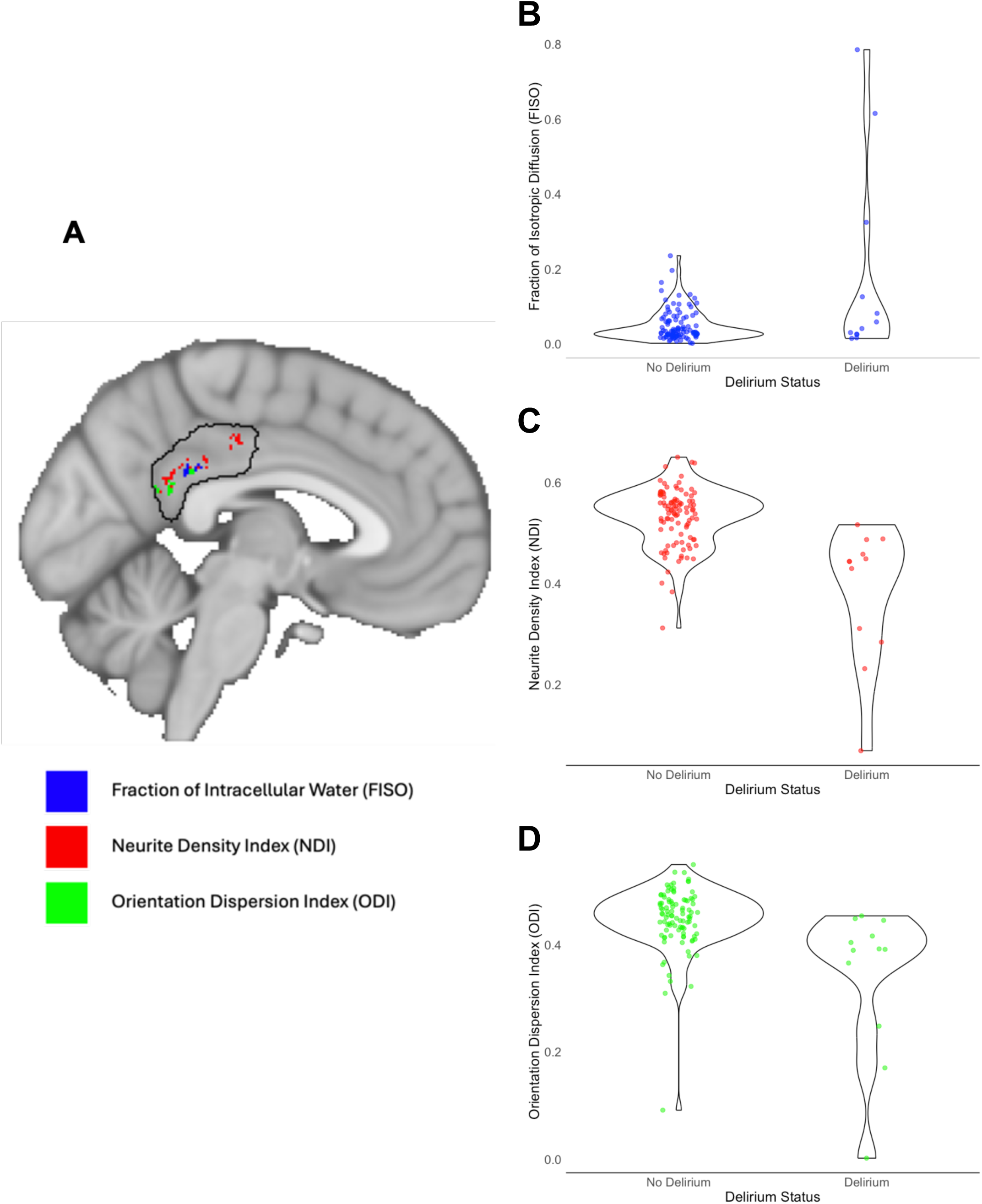
PCC region of interest constrained diffusion metric analysis, from preoperative MRI scans. (A) Overlay of significant (p < 0.05) voxels for all three diffusion metrics. (B-D) Violin plots displaying the distribution of each diffusion metric by postoperative delirium status: (B) Fraction of isotropic diffusion (FISO), (C) Neurite Density Index (NDI), and (D) Orientation Dispersion Index (ODI).

**Table 2:**
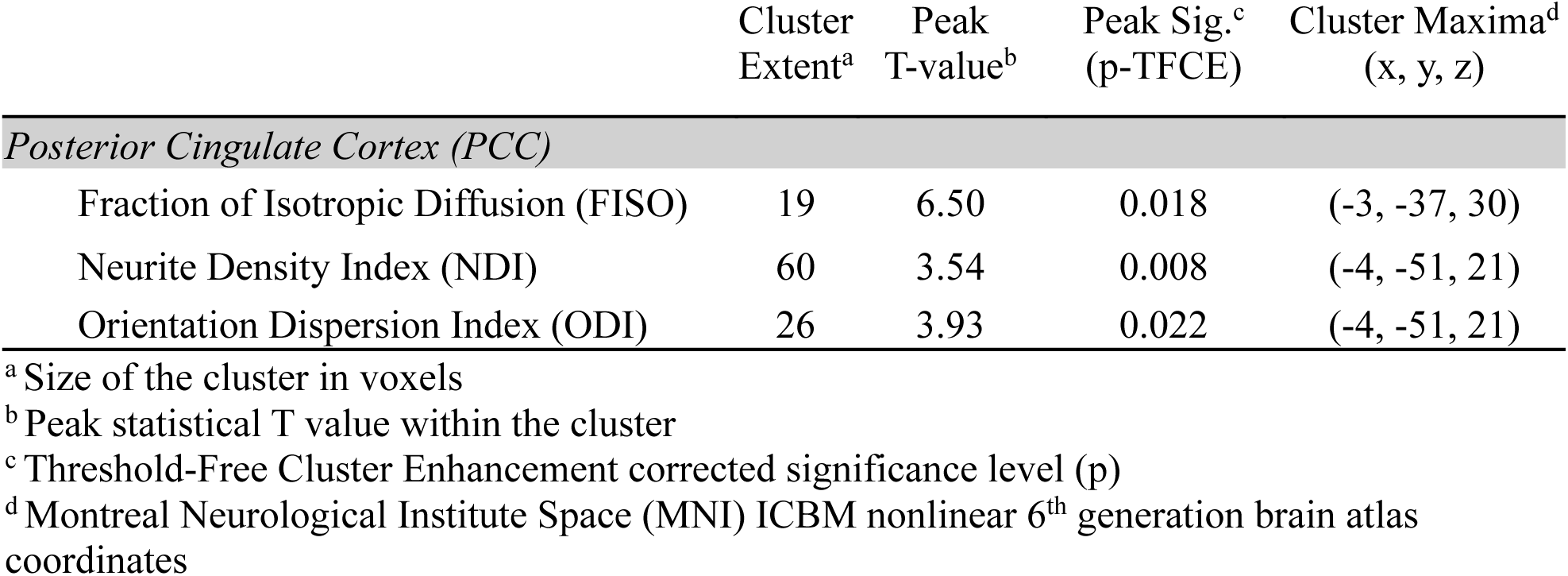
NODDI peak and cluster values within the PCC region of interest, from preoperative MRI scans, that differed among patients who later developed (vs those who did not develop) postoperative delirium.

### Dorsal Posterior Cingulate Cortex Microstructure Relates to Preoperative Cognition

Next, we explored relationships between these significant baseline diffusion metrics and cognitive function domains across the entire sample (Figure 3). Higher NDI and ODI values within this dorsal PCC subcluster were significantly associated with better performance on the attention and concentration factor (NDI: r = 0.222, p = 0.021; ODI: r = 0.270, p = 0.005, respectively; see Figure 3). Yet they were not significantly associated with structured auditory-verbal memory, unstructured auditory-verbal memory, visual memory, or information processing speed/executive functioning (Figure 3), nor were they associated with overall cognitive index (p > 0.05 for each, Figure 4).

**Figure 3:**
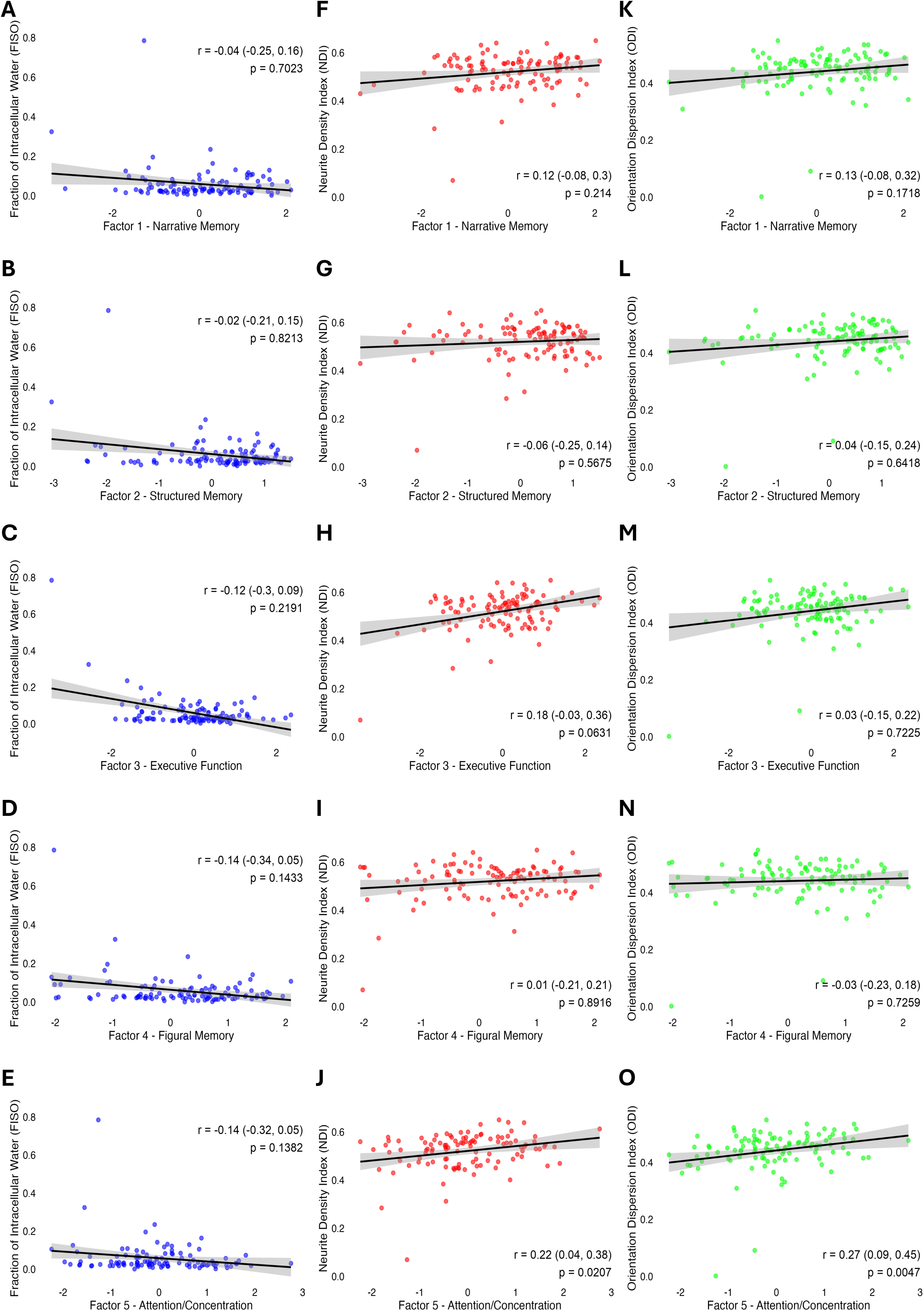
Scatter plots illustrating the relationships between diffusion metrics and cognitive factors. Spearman’s r, 95% confidence intervals and p values are provided for each plot. (A-E) Fraction of isotropic diffusion (FISO), (F-J) Neurite Density Index (NDI), (K-O) Orientation Dispersion Index (ODI). Each NODDI metric is subdivided by row for each of the following cognitive factors: Factor 1 - Narrative Memory, Factor 2 - Structured Memory, Factor 3 - Executive Function, Factor 4 - Figural Memory, and Factor 5 - Attention/Concentration. Significant associations were identified via Spearman’s (r) between both NDI (J) and ODI (O) and Factor 5 - Attention/Concentration (p < 0.05), respectively. All other associations were not significant (p > 0.05).

**Figure 4:**
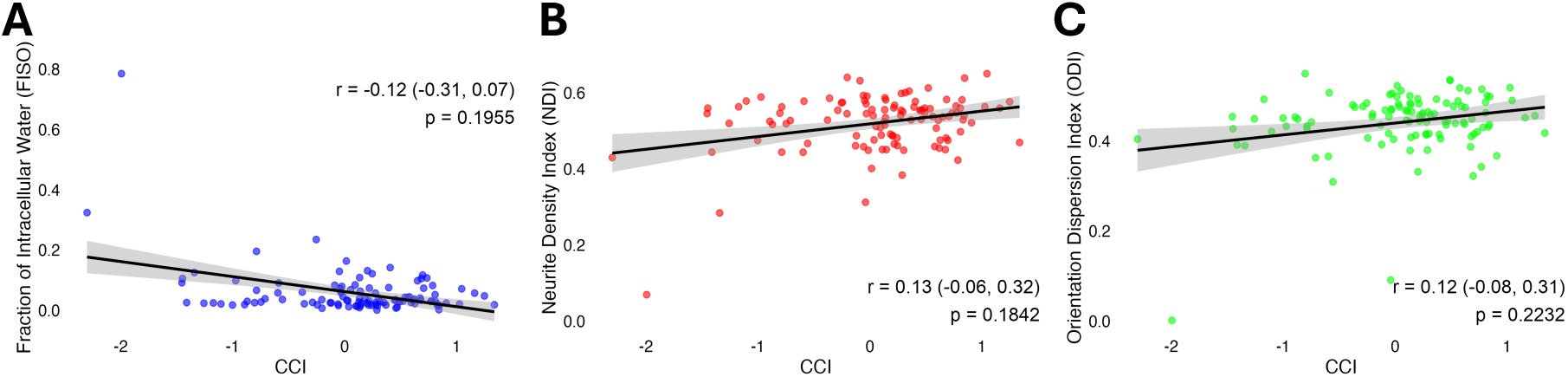
Scatter plots illustrating the relationships between diffusion metrics and global cognitive function (Continuous Cognitive Index, or CCI). Spearman’s r, 95% confidence intervals, and p values are provided for each plot. No significant relationships (p > 0.05) were identified via Spearman’s (r) between any of the three NODDI metric (A) Fraction of Isotropic Diffusion (FISO), (B) Neurite Density Index (NDI), (C) Orientation Dispersion Index (ODI) and CCI.

Next, we considered the possibility that the relationship between these diffusion metrics and attention/concentration might reflect potential confounding from age, education or brain atrophy. Importantly, the multivariable regression analysis revealed that the associations of both NDI and ODI, but not FISO with attention/concentration remained significant after adjustment (FISO: β̂ = -0.277 (95% CI: -0.599 - 0.046), p = 0.092; NDI: β̂ = 0.292 (95% CI: 0.095 - 0.489), p = 0.004; ODI: β̂ = 0.290 (95% CI: 0.101 - 0.479), p = 0.003). Bootstrapped confidence intervals for the regression coefficients indicated that NDI was a significant predictor in 84% of samples (β̂ 95% CI: 0.131 – 0.473) and ODI was a significant predictor in 90% of samples (β̂ CI: 0.063 – 0.430).

### Dorsal Posterior Cingulate Cortex Microstructure Does Not Relate to Amyloid Status

Lastly, given the known accumulation of Aβ in the PCC in early AD,^17^ we evaluated the potential influence of Aβ pathology, as assessed by CSF Aβ levels, on preoperative/baseline diffusion metrics across subjects. Multivariable linear regression adjusted for age, education and global brain atrophy, revealed no significant relationship between Aβ positivity (n = 22 positive) and diffusion metrics (FISO: β̂ = 0.000 (95% CI: -0.003 - 0.004),p = 0.797; NDI: β̂ = -0.015 (95% CI: -0.005 - 0.002), p = 0.357; ODI: β̂ = 0.001 (95% CI: -0.002 - 0.004), p = 0.452). Furthermore, in analysis of numeric Aβ levels from INTUIT subjects (n = 53), there was no significant correlation between CSF Aβ levels and any of the diffusion metrics in the dorsal PCC subcluster (FISO Spearman’s r = -0.021, 95% CI: -0.332 - 0.269, p = 0.881; NDI Spearman’s r = 0.008, 95% CI: -0.264 - 0.269, p = 0.955; ODI Spearman’s r = -0.242, 95% CI: -0.485 - 0.035, p = 0.081) (Figure 5). Similarly, these patterns did not change after multivariable adjustment.

**Figure 5:**
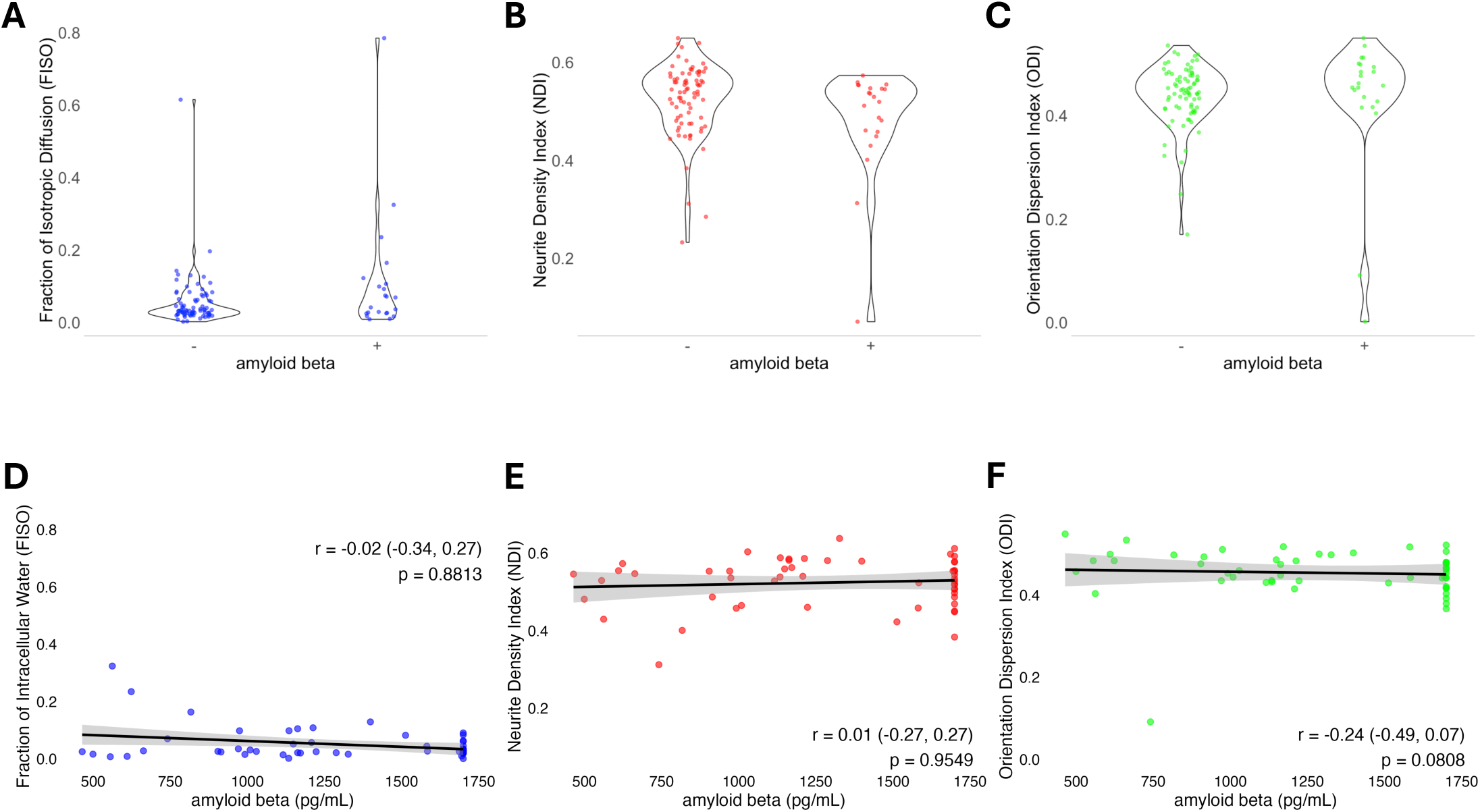
Violin and Scatter plots illustrating the relationships between diffusion metrics and cerebrospinal fluid Aβ. **A-C)** Diffusion metrics by Aβ42 positivity status. No significant differences were found in diffusion metrics between those with versus those without positive CSF levels of Aβ. **D-F)** Scatter plots for diffusion metrics by numeric levels of Aβ. Spearman’s r, 95% confidence intervals and p values are provided for each plot. No significant relationships (p > 0.05) were identified via Spearman’s (r) between the diffusion metrics **(A, D)** Fraction of isotropic diffusion (FISO), **(B, E)** Neurite Density Index (NDI), **(C, F)** or Orientation Dispersion Index (ODI) with CSF Aβ levels.

## Discussion

Here we show for the first time that older non-cardiac/non-intracranial surgery patients who developed (versus those who did not develop) postoperative delirium had higher FISO, as well as lower NDI and ODI within the dorsal PCC before surgery. Importantly, these diffusion metrics were not significantly related to CSF measures of Aβ, suggesting that these microstructural markers may represent a distinct vulnerability beyond CSF AD-related Aβ neuropathology. Furthermore, in multivariable analyses controlling for demographic and structural brain factors, these alterations in baseline/preoperative PCC microstructure remained significantly associated with preoperative cognition, specifically the attention/concentration cognitive domain score. This demonstrates that the relationship between these preoperative diffusion metrics and preoperative attention/concentration is independent of (and not driven by) age, education, or global brain atrophy. Taken together, these findings identify preoperative microstructural gray matter differences within the PCC that are associated with preoperative attention/concentration deficits, and which are associated with (and may thus underlie vulnerability to) the development of delirium after surgery.

Our finding of greater preoperative differences in NDI and ODI in subjects who went on to develop delirium contrasts with NODDI studies of mild cognitive impairment and AD, which have reported greater differences between groups (i.e., MCI/AD versus cognitively normal) within multiple brain regions in FISO (compared to NDI or ODI).^52–55^ Our data indicates delirium is associated with alterations in axonal/dendritic density (NDI) and diffusion around these processes (ODI), whereas other studies show that the ADRD spectrum is more commonly associated with increases in extracellular free water that can diffuse in all three dimensions (i.e., fluid filled space that is not immediately next to neuronal processes). In essence, these findings suggest that there may be distinct types of microstructural brain differences within different brain regions in patients that go on to develop postoperative delirium versus those seen in the ADRD spectrum.

The microstructural differences within the PCC in patients that later developed postoperative delirium are interesting, because the PCC is a key region of the brain’s default mode network (DMN),^56^ and is known to play a role in network switching and attentional control.^57,58^ Functional MRI studies have shown that the PCC is highly active during rest and plays a central role in internally directed attention, such as during autobiographical memory retrieval and self-referential thought.^57^ Furthermore, studies demonstrate that the DMN, and in particular the PCC, is anticorrelated with both the cognitive control network^59^ and the dorsal attention network,^57^ each of which are active during the performance of attentionally demanding tasks. One interpretation of these observations is that suppression of PCC activity is critical for external attentional focus and concentration.^59,60^ Moreover, studies have shown that disruptions to the normal anticorrelation between the PCC and task-positive regions were associated with postoperative delirium.^9,11^ Therefore, latent PCC microstructural damage (i.e., in patients without neurologic diagnoses) may indeed disrupt functional activity in this brain region, which in turn could lead to deficits in attentional control, a key feature of postoperative delirium.

The DMN, including the PCC, has also been identified as the earliest area in which Aβ pathology develops in AD.^17^ Parker et al.^54^ found significant changes in diffusion metrics within temporal and parietal lobe gray matter in patients with early-onset AD, and these microstructural changes were associated with the magnitude of cognitive impairments. This study also found that patients with early-onset AD had diffusion metric changes in the precentral gyrus of the frontal lobe, a region associated with AD-related pathology but typically spared from gross volume and cortical thickness changes in AD.^54^ Given the relationship between NODDI changes in regions associated with AD, and that the PCC is an early site of Aβ deposition, we explored the potential relationship between diffusion metrics within the PCC and CSF Aβ levels, and found no significant association. Although speculative, this finding might suggest that our novel biomarker of PCC microstructural vulnerability represents a valuable predictor of the subsequent development of delirium, though the relationship between diffusion metrics and AD neuropathologies in this population requires further study.

The debate over whether postoperative delirium represents an unmasking of latent preoperative vulnerabilities or is itself injurious remains unresolved, though these possibilities are likely not mutually exclusive. In our data, all three diffusion metrics (i.e., FISO, NDI, and ODI) differed significantly before surgery, between those who later developed postoperative delirium and those who did not. Similarly, by using more traditional diffusion MRI measures in patients who subsequently developed (versus those who did not develop) postoperative delirium, Cavallari et al.^61^ identified diffuse preoperative white matter microstructural differences in parietal, temporal, and occipital cortices, as well as in regions such as the cerebellum, thalamus, and cingulum. These findings, along with ours, strongly support the hypothesis that preexisting/preoperative latent brain abnormalities—both in gray and white matter—predispose certain patients to postoperative delirium. Yet, this does not rule out the contribution of additional perioperative precipitating factors to delirium risk, which likely act in concert with predisposing factors (such as preoperative localized structural abnormalities within the brain), to ultimately lead to postoperative delirium.^7,62^ Moreover, while we do not find large effect sizes for changes in diffusion metrics six weeks post-surgery, a longitudinal study by Cavallari et al.^63^ revealed greater changes in frontal, temporal, and parietal white matter microstructure between baseline and one-year follow-up in patients who developed postoperative delirium. Thus, an important goal for future studies is to understand how preoperative neurologic abnormalities (even if subclinical and undiagnosed) interact with surgical stress, inflammation, and other precipitating factors to result in the clinical syndrome of delirium and how delirium may alter microstructural integrity during the first year after surgery.

These findings should be interpreted with caution. First, because of the exploratory nature of our correlation analyses, we did not correct for multiple comparisons in Spearman correlation or linear regression analyses. Second, although this study cohort had over 100 patients, the number who developed postoperative delirium was small (n = 12); thus, this data set is not suitable for more in-depth analyses including mediation. However, the delirium rate (∼11%) in this cohort is within the expected range for non-cardiac/non-intracranial surgery patients aged ≥ 60 years. Further, this study’s sample size (N=111 with diffusion data before and 6 weeks after surgery) is similar to that of the Cavallari et al^63^ study, in which 113 subjects had both baseline and one-year follow-up MRI scans, and larger than the Choi et al.^9^ study in which 14 patients who developed postoperative delirium had MRI at both timepoints. Participants in these two other studies were older on average (76 ± 4 and 73.6 ± 9.4 years, respectively) than those in this study; however, the relatively younger cohort (i.e., mean age 68.32 ± 5.72 years) in this study enhances the generalizability of these findings to the larger population of older surgical patients. Third, to examine differences between patients who later developed (vs those who did not develop) postoperative delirium, we applied a region of interest-specific restriction to the PCC. Larger future, higher-powered diffusion imaging studies should confirm these findings within this PCC region of interest and conduct additional brain-wide analyses. Indeed, a much larger sample size would likely be required for detecting PCC differences at a whole-brain level using threshold-free cluster enhancement (TFCE). Additionally, post-mortem histology, immunohistochemistry, spatial transcriptomics, and electron microscopy could be used to identify the molecular and cellular underpinnings of diffusion abnormalities within the dorsal PCC of patients who developed postoperative delirium. This multidisciplinary approach would enhance confidence in the PCC-specific findings presented here and could explore broader brain-wide associations with delirium. Fourth, while we found differences in all three diffusion metrics within the dorsal PCC between these groups before surgery, we found that only FISO values (but not NDI or ODI metrics) within the dorsal PCC remained significantly different among patients with (vs those without) postoperative delirium at 6 weeks after surgery. One potential explanation for the inconsistency of significant findings between time points can likely be attributed to increased data variability due to the heterogeneity of surgical procedures and postoperative patient trajectories￼ This is supported by additional analyses where the p-value threshold was relaxed to a statistical trend (p < 0.08) at the 6-week postoperative timepoint, between-group differences in all three diffusion metrics within the dorsal PCC emerged between groups, similar to our preoperative results.

Nonetheless, this study also had several strengths: First, we used validated delirium assessments, and our delirium assessors were all trained by a neuropsychologist. Furthermore, the majority of patients whose data is included in this work were from the INTUIT study, in which delirium assessments were conducted twice daily until discharge. Together, these measures decrease subjectivity and increase sensitivity to detect postoperative delirium. Second, all patients whose data is included in this work underwent detailed neuropsychological testing before surgery, providing a comprehensive measure of preoperative cognitive function. This allowed us to relate preoperative neuroimaging metrics with cognitive function before the development of postoperative delirium (and any post-delirium cognitive impairments), a novel feature of this work. Third, patients in this study underwent research lumbar punctures allowing us to measure preoperative CSF Aβ levels and to determine their association with preoperative diffusion imaging metrics in the PCC. Fourth, this is the first study to use NODDI diffusion metrics to identify preoperative gray matter microstructural abnormalities in patients who later developed postoperative delirium, and our results suggest that gray matter abnormalities in the PCC represent a novel neuroanatomic locus that may predispose some patients to postoperative delirium. Thus, the findings presented here, together with prior work,^9–14^ implicate the PCC as a key hub of functional and microstructural abnormalities associated with both preoperative cognitive impairment and the subsequent development of postoperative delirium in older surgical patients.

## Data availability

Data that support the findings of this study are available from the senior author upon reasonable request once the necessary data use agreements are signed.

## Acknowledgements

We thank Drs. Allen Song and Todd Harshbarger for assistance with MRI scans, and Susan Music, Lamont Conyers, and Jennifer Graves for helping ensure that all participants were safe to undergo 3T MRI scans. We also thank the older surgical patients who consented and took the time to participate in this work, around the same time as they were undergoing a major surgery under general anesthesia.

