## Supplementary Materials for "Predilection for Perplexion: Preoperative microstructural damage is linked to postoperative delirium"

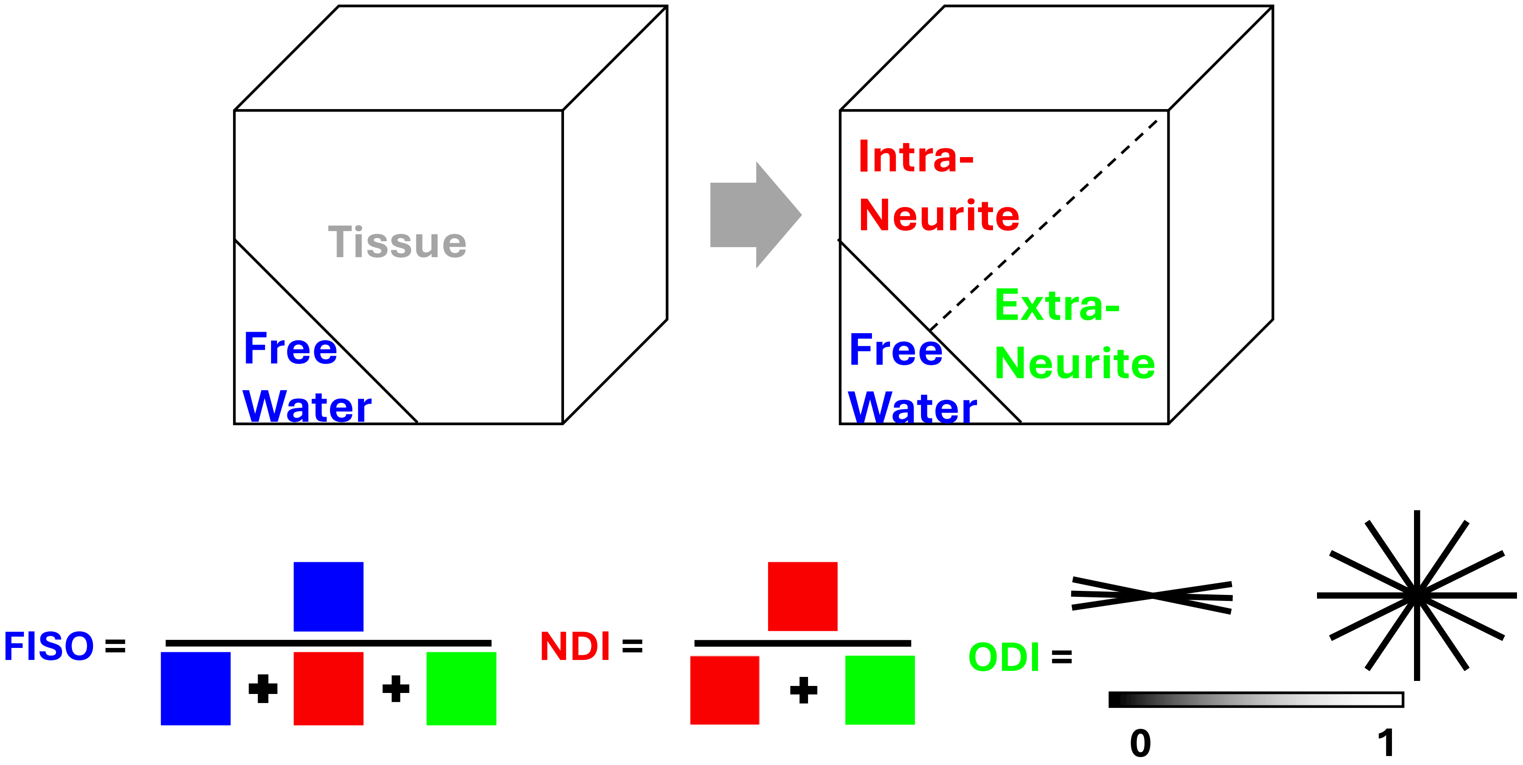


**Supplementary Figure 1:** Schematic representation of NODDI (Neurite Orientation Dispersion and Density Imaging) metric calculations. The model separates diffusion MRI signals into isotropic diffusion (i.e., free water) and the neurite compartment, from which intracellular and extracellular diffusion are derived. From these components, three diffusion metrics are derived:

- FISO (Fractional Isotropic Volume): The proportion of free (i.e., extracellular) water, derived from the isotropic compartment of the model.
- NDI (Neurite Density Index): Reflects the fraction of intracellular diffusion, representing neurite density, estimated from the intracellular compartment.
- ODI (Orientation Dispersion Index): Measures the angular variability of neurite orientations, derived from the extracellular compartment through the orientation dispersion model.

Adapted from http://mig.cs.ucl.ac.uk/index.php?n=Tutorial.NODDImatlab

**Supplementary Table 1:** Cognitive domain factors and individual test scores

| | |
|---|---|
| **Factor 1 Unstructured (Narrative) Memory Domain (z)** | **0.10 (1.06)** |
| RMT Immediate Recall – Gist Score | 7.37 (1.59) |
| RMT Immediate Recall – Verbatim Score | 11.3 (3.21) |
| RMT Delayed Recall – Gist Score | 6.93 (1.87) |
| RMT Delayed Recall – Verbatim Score | 9.73 (3.4) |
| **Factor 2 Structured (Word List) Memory Domain (z)** | **0.17 (0.90)** |
| HVLT-R Immediate Recall Total Score (Sum of Trials 1-3) | 6.1 (23.9) |
| HVLT-R Delayed Recall Score | 7.88 (3.66) |
| HVLT-R Delayed Recognition Discrimination Index Score | 10.4 (1.79) |
| **Factor 3 Processing Speed / Executive Function Domain (z)** | **0.05 (0.94)** |
| WAIS-R Digit Symbol Substitution Subtest Score | 43.2 (11.3) |
| Trail Making Test – Part A completion time (sec.) * | 38.9 (29.9) |
| Trail Making Test – Part B completion time (sec.) * | 115 (74.2) |
| **Factor 4 Visual (Figural) Memory Domain (z)** | **0.16 (0.97)** |
| WMS-R Visual Reproduction Immediate Recall Score | 6.68 (2.73) |
| WMS-R Visual Reproduction Delayed Recall Score | 6.16 (2.89) |
| **Factor 5 Attention & Concentration Domain (z)** | **0.06 (1.03)** |
| WAIS-R Digit Span Subtest – Forward Span Score | 7.46 (2.18) |
| WAIS-R Digit Span Subtest – Backward Span Score | 6.37 (2.28) |

All data are presented as mean (SD)
RMT – Randt Memory Test
HVLT-R – Hopkins Verbal Learning Test – Revised
*To include Trail Making Test scores in the Factor 3 scores along with WAIS-R Digit Symbol Substitution (in which higher scores are better), we transformed the Trail Making Test scores listed above by subtracting performance in seconds from the maximum time (300 sec.) such that higher trails scores (which measure time remaining after task completions) indicated better performance.

**Supplementary Table 2:** Perioperative Factors

| | **Overall** | **No Delirium** | **Delirium** |
|---|---|---|---|
| **Surgical Service (%)** | | | |
| Cardiothoracic | 14 (12.6) | 13 (13.1) | 1 (8.3) |
| General Surgery | 29 (26.1) | 24 (24.2) | 5 (41.7) |
| Gynecology | 5 (4.5) | 5 (5.1) | 0 (0) |
| Orthopedics | 29 (26.1) | 26 (26.3) | 3 (25) |
| Otolaryngology Head and Neck | 3 (2.7) | 2 (2) | 1 (8.3) |
| Plastic Surgery | 3 (2.7) | 3 (3) | 0 (0) |
| Urology | 28 (25.2) | 26 (26.3) | 2 (16.7) |
| **Surgical Duration (minutes) [Q1, Q3]** | 140 [104, 189] | 136 [99, 178] | 207 [146, 326] |
| **Propofol (mg)** | 558.73 (773.52) | 530.63 (715) | 790.5 (1167.55) |
| **Ketamine (mg)** | 12.58 (23.85) | 12.84 (24.39) | 10.42 (19.59) |
| **Dexmedetomidine (mcg)** | 4.18 (15.00) | 4.41 (15.75) | 2.33 (6.02) |
| **Intraoperative Opioid Dose (OME)** | 16.52 (11.73) | 16.41 (11.82) | 17.35 (11.47) |
| **Muscle Relaxants*** | 60.37 (49.52) | 59.12 (48.67) | 70.67 (57.34) |
| **aaMAC**** | 0.80 (0.20) | 0.8 (0.2) | 0.86 (0.18) |
| **Estimated Blood Loss (mL) [Q1, Q3]** | 75 [10, 200] | 50 [10, 175] | 175 [76.3, 325] |

Data are presented as mean (SD) or median [Q1, Q3]
OME –oral morphine equivalents
*In Rocuronium dose equivalents[51]
**aaMAC – age adjusted end tidal mean alveolar concentration of anesthetic gas.

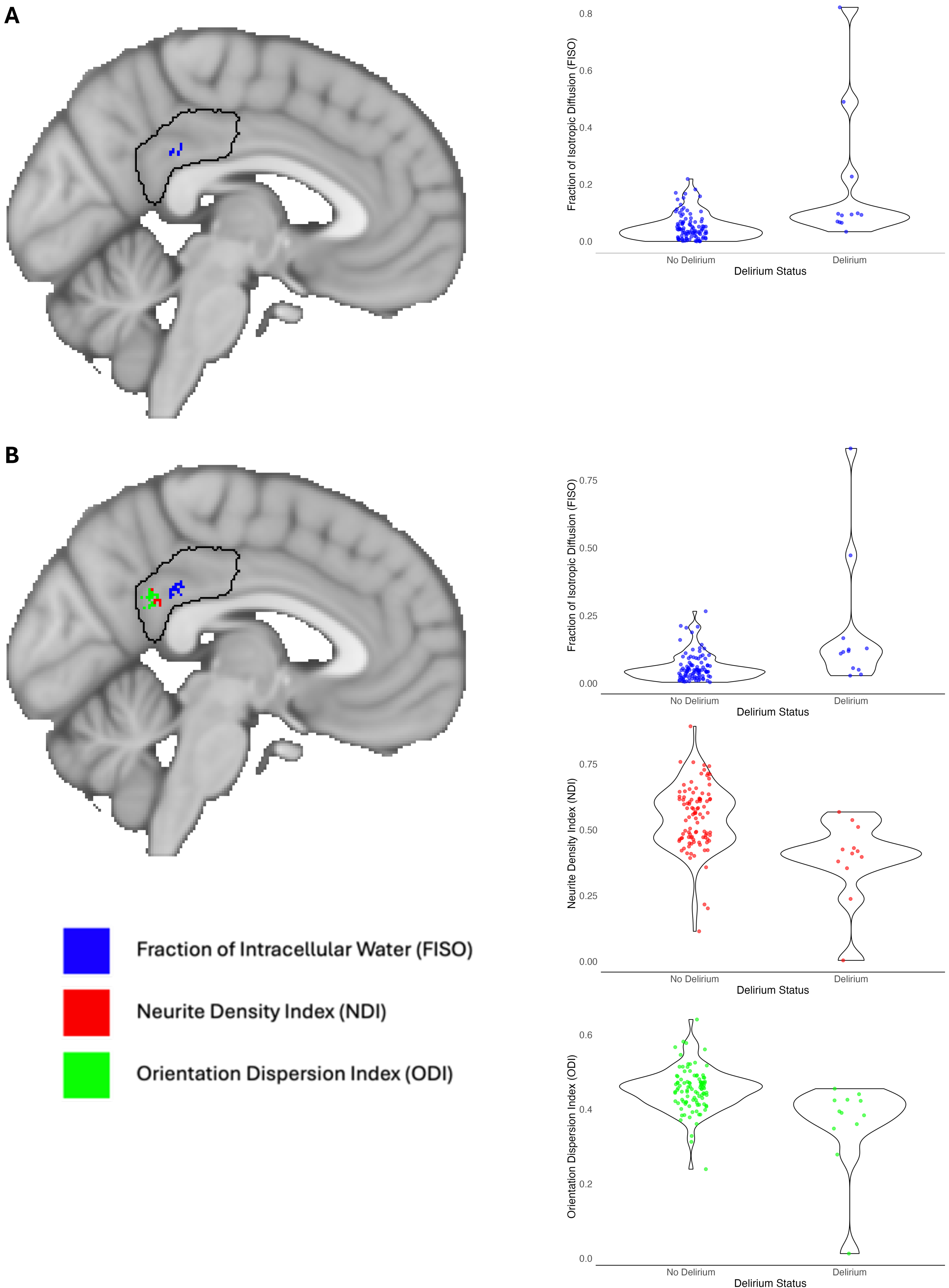
A
B
Fraction of Isotropic Diffusion (FISO)
0.8
0.6
0.4
0.2
0.0
No Delirium
Delirium
Delirium Status
Fraction of Isotropic Diffusion (FISO)
0.75
0.50
0.25
0.00
No Delirium
Delirium
Delirium Status
Neurite Density Index (NDI)
0.75
0.50
0.25
0.00
No Delirium
Delirium
Delirium Status
Orientation Dispersion Index (ODI)
0.6
0.4
0.2
0.0
No Delirium
Delirium
Delirium Status
Fraction of Intracellular Water (FISO)
Neurite Density Index (NDI)
Orientation Dispersion Index (ODI)

**Supplementary Figure 2:** PCC constrained diffusion metric analysis at 6 weeks after surgery. (A) Overlay of significant ($p < 0.05$) voxels for FISO and corresponding violin plot. (B) Overlay of suprathreshold voxels when relaxed to statistical trend ($P < 0.08$) and corresponding violin plots for each diffusion metric.

**Supplementary Table 3:** NODDI peak and cluster values within the PCC region of interest, from 6-week postoperative MRI scans, that differed among patients who later developed (vs those who did not develop) postoperative delirium.

| | Cluster Extent[a] | Peak T-value[b] | Peak Sig.[c] (p-TFCE) | Cluster Maxima[d] (x, y, z) |
|---|---|---|---|---|
| *Threshold p < 0.05* | | | | |
| Fraction of Isotropic Diffusion (FISO) | 5 | 3.56 | 0.022 | (-3, -39, 30) |
| *Threshold p < 0.08* | | | | |
| Fraction of Isotropic Diffusion (FISO) | 18 | 5 | 0.022 | (-3, -41, 31) |
| Neurite Density Index (NDI) | 10 | 2.64 | 0.053 | (-3, -47, 27) |
| Orientation Dispersion Index (ODI) | 23 | 3.01 | 0.043 | (-3, -49, 25) |

[a] Size of the cluster in voxels

[b] Peak statistical T value within the cluster

[c] Threshold-Free Cluster Enhancement corrected significance level (p)

[d] Montreal Neurological Institute Space (MNI) ICBM nonlinear 6th generation brain atlas coordinates
